# Association of preference and frequency of teleworking with work functioning impairment: a nationwide cross-sectional study of Japanese full-time employees

**DOI:** 10.1101/2021.11.04.21265947

**Authors:** Satoshi Yamashita, Tomohiro Ishimaru, Tomohisa Nagata, Seiichiro Tateishi, Ayako Hino, Mayumi Tsuji, Kazunori Ikegami, Keiji Muramatsu, Yoshihisa Fujino, the CORoNaWork Project

## Abstract

**Objective:** We examined whether teleworking preference and frequency were associated with work functioning impairment.

**Methods:** This online cross-sectional study was conducted using a self-administered questionnaire among 27,036 full-time Japanese workers. The Work Functioning Impairment Scale was used to measure work functioning impairment, and we performed multilevel logistic regression analysis.

**Results:** Higher odds ratios for work functioning impairment were observed among employees who preferred to telework compared with those who preferred working in the workplace. A similar trend was observed among employees who teleworked 4 or more days a week compared with those who almost never teleworked. When teleworking preference and frequency were adjusted, only teleworking preference was associated with work functioning impairment.

**Conclusions:** A preference for teleworking was associated with work functioning impairment and one factor that increased the teleworking frequency.

## Introduction

Teleworking has increased among companies worldwide during the COVID-19 pandemic to prevent contact among workers and limit the spread of infection. ^1,2^ Telework involves working at a location other than a central office or workplace, such as at home. Individuals work separately from other employees during working time and use information and communications technology to interact with coworkers, as needed.^3^ A recent survey in Japan revealed that 26.7% of employees were teleworking as of March 2020, the beginning of the COVID-19 pandemic, and this percentage increased to 52.7% as of May 2020 during the first the state of emergency declaration by the Japanese government.^4^ A previous survey in Japan showed that the percentage of teleworkers peaked in May 2020 at more than 50%; however, after the first state of emergency declaration period, the percentage of teleworkers had decreased to approximately 30% by June 2020.^5^ To not only prevent infection but also maintain business continuity, some workers combine teleworking with going to their office or workplace.^6^

Teleworking is considered to have both positive and negative effects on workers’ health. Teleworking is characterized by the absence of commuting time and flexibility regarding working location and time. However, teleworking also involves a lack of supervision by superiors, sitting and working at home for long periods, and little communication with colleagues.^3,7–9^ The positive effects of teleworking on workers include the reduction in commuting time, which allows for more sleep, fewer interruptions from colleagues at work, and greater flexibility in how employees use their time to improve their work–life balance.^3,7,8,10^ However, the negative effects of teleworking on workers include musculoskeletal disorders owing to an inappropriate work environment, excessive weight gain because of physical inactivity, increased risk of cardiovascular disease and diabetes, and adverse effects on mental health owing to limited opportunities to interact with colleagues and superiors.^3,9^ Teleworking affects the ease of work and performance through its impact on workers’ health.

The negative effects of teleworking may cause work functioning impairment. Work functioning impairment is loss of the ability to perform a work task as a result of presenteeism, which is defined as “reduced performance at work, besides illness.”^11,12^ A teleworking system not only permits employees to work flexibly but it also enables them to work, even when they have a health problem, such as fatigue or depression.^8,13^ Previous studies have shown that teleworking increases sedentary behavior and that sedentary behavior leads to presenteeism.^8,14^ The introduction of teleworking may promote presenteeism and working with some degree of work functioning impairment.

It is not known how teleworking affects work functioning impairment. In particular, the rapid introduction of teleworking during the early part of the COVID-19 pandemic may have created a mismatch between workers’ preferences and teleworking practices. This rapid introduction of teleworking differed from teleworking before the COVID-19 pandemic because workers could not choose to work remotely at will. The option of teleworking was previously selected according to workers’ preferences; however, teleworking during the COVID-19 pandemic was imposed owing to safety and social requirements, regardless of workers’ preferences. Imposing shift work on workers who do not prefer shift work is associated with distress and burnout.^15^ However, the impact of a preference for teleworking among workers is unclear. In this study, we examined how teleworking preferences and frequency are associated with work functioning impairment.

## Methods

### Study design and participants

This was a cross-sectional study using online questionnaire survey data from the Collaborative Online Research on Novel-coronavirus and Work study (CORoNaWork study). The questionnaires were sent by e-mail to target populations who were preregistered with Cross Marketing Inc. (Tokyo, Japan) and monitors who responded so as to participate in the survey. The data collection period was from December 22 to 26, 2020. To recruit participants, 605,381 people were emailed a request to participate in the survey; 55,045 people were enrolled as monitors, and 33,087 people matched the survey criteria. The survey was completed by 33,087 Japanese full-time workers between the ages of 20 to 65 years. After excluding 6,051 surveys with invalid responses, 27,036 people were included in the analysis (Figure 1). Invalid responses were those from surveys completed in less than 6 minutes; participants with weight less than 30 kg or height less than 140 cm; inconsistent responses; and wrong answers to the following question: Select the third largest number out of the following five numbers (231, 245, 323, 252, 312, and the correct answer is 252). Stratified sampling was conducted by region (five regions according to the regional COVID-19 incidence level), sex (male/female), and job type (office worker/non-office worker). National statistics were used to obtain the regional COVID-19 incidence level during the study period.^16^ We classified this into five levels according to the cumulative incidence of COVID-19 in each prefecture. Thus, respondents were categorized into 20 units sampled by region, sex, and job type. Each unit had an equal number of 1650 respondents. Recruitment continued until the required number was reached in each unit. Details of the sample selection have been described elsewhere.^17^ This study was conducted with the approval of the Ethics Committee of the University of Occupational and Environmental Health, Japan (approval no. R2-079). Informed consent was obtained via a form on the survey website.

**Figure 1.**
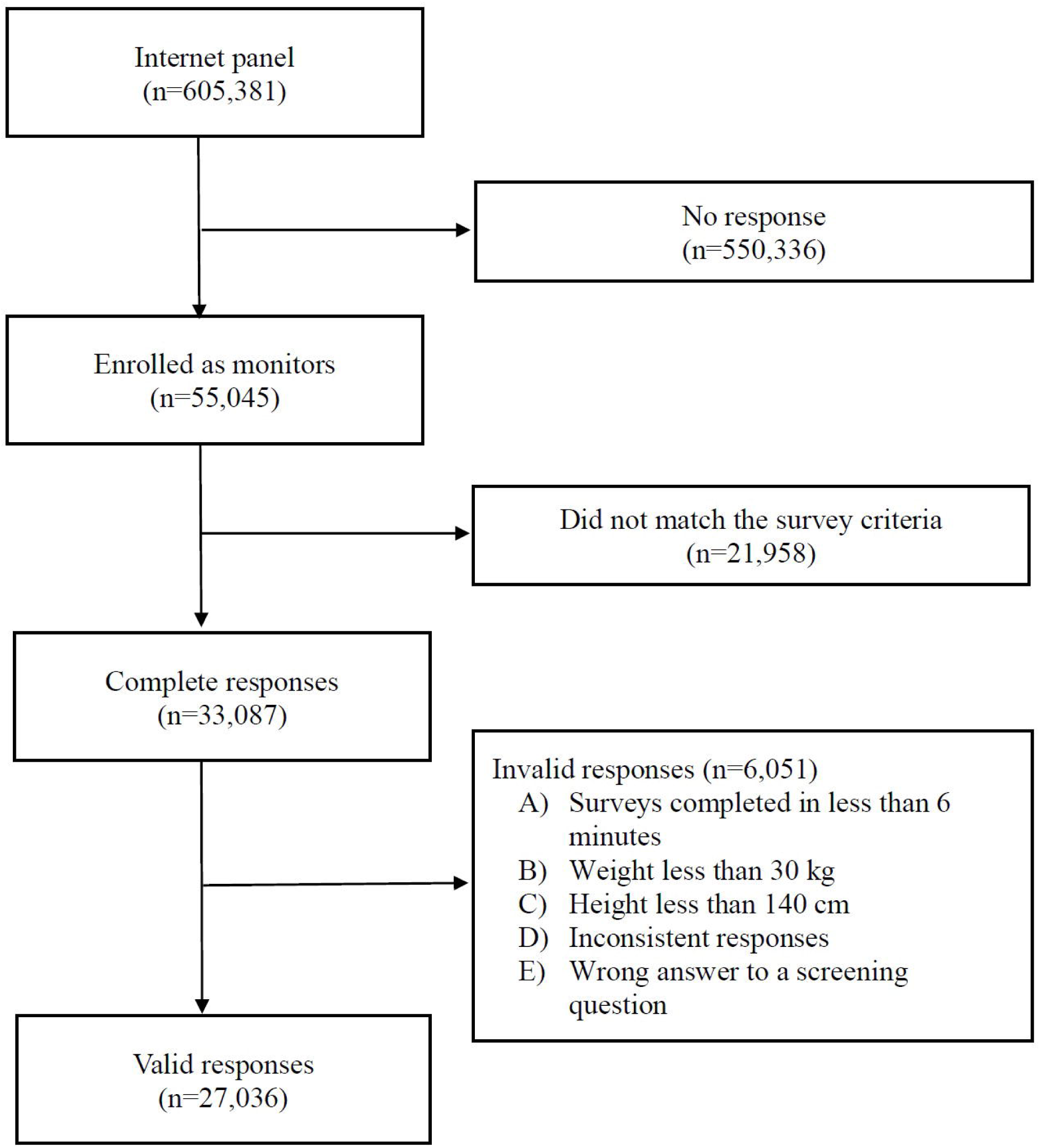
Flow diagram of the study population

### Outcome

The Work Functioning Impairment Scale (WFun) was used as a measure of work functioning impairment. The WFun is a self-administered measure of work functioning impairment validated and developed according to the Consensus-based Standards for the Selection of Health Measurement Instruments and based on the Rasch model.^18^ The instrument comprises seven items, with five response options. The total of the seven items is the overall score, which ranges from 7 to 35 points, with higher scores indicating higher work functioning impairment. We used the 6-item version of the WFun. This simplified version is also based on the Rasch model and is converted to the same score as in the original measurement instrument by multiplying by 1.17 (7–35 points).^19,20^ We defined work functioning impairment as a WFun score of 21 or higher, in line with previous studies.^21^

### Explanatory variables

Teleworking frequency and teleworking preferences were included as explanatory variables. Participants responded to the following question about teleworking preferences: “How do you feel about teleworking?” with the following options: a) I would like to telework as much as possible; b) I prefer teleworking; c) I don’t mind either; d) I prefer regular work; and e) I prefer regular work as much as possible. Participants responded to the following question regarding teleworking frequency: “Do you work from home?” with the following response options: a) 4 or more days a week; b) 2 or 3 days a week; c) 1 day a week; d) more than once a month and less than once a week; and e) almost never.

### Covariates

On the basis of relevant studies, the following socioeconomic and lifestyle factors were selected as covariates: sex,^22,23^ age,^22–24^ marital status^22^ (married, divorced/widowed, and unmarried), annual equivalent household income,^24^ education^23^ (junior high school, high school, vocational school, junior college, university, and graduate school), smoking,^22^ alcohol consumption,^22^ job type^14,23,25^ (mainly desk work, mainly involving interpersonal communication, mainly physical work), number of employees,^4^ and COVID-19 incidence rate in the month prior to the survey.^4^

### Statistical analysis

A WFun score of 21 was used as the cutoff value. Scores above 21 were dichotomized as work functioning impairment and scores below 21 as no work functioning impairment. For teleworking frequency, “once a week” was included in “more than once a month and less than once a week” owing to the small number of respondents. Multilevel logistic regression analysis was performed to assess the relationship of teleworking preference and frequency with WFun score. The hierarchical structure nested in the prefecture of residence according to the cumulative incidence of COVID-19 in each prefecture was adjusted to account for the effects of intraclass correlation. The associations between teleworking and work functioning impairment were evaluated using three models: an age- and sex-adjusted model, Model 1, and Model 2. In Model 1, we adjusted for sex, age, marital status, annual equivalent household income, education, smoking, alcohol consumption, job type, number of employees, and COVID-19 incidence rate. Model 2 was adjusted for teleworking preferences and teleworking frequency, in addition to those factors adjusted in Model 1. All statistical analyses were conducted using Stata/SE 16.1 (StataCorp LLC, College Station, TX, USA). A p-value less than 0.05 was considered statistically significant.

## Results

The characteristics of respondents are shown in Table 1. Of 27,036 respondents, approximately half were men (51.1%) and office workers (49.8%). Approximately three-quarters (72.9%) had a technical college, junior college, university, or graduate degree. Most respondents (78.7%) reported nearly no teleworking; 10.3% of respondents reported very frequent (4 or more days a week) teleworking. Respondents who reported nearly no teleworking had the lowest percentage (22.1%) of high WFun scores. There were no missing data because the survey system was designed to ensure that all questions were answered.

**Table 1.** Participant characteristics

|  | Teleworking frequency |  |  |  |  |  |  |  |
| --- | --- | --- | --- | --- | --- | --- | --- | --- |
|  | 4 or more days a week<br>(n=2,790) |  | 2 or 3 days a week<br>(n=1,477) |  | More than once a month and less than once a week<br>(n=1,493) |  | Almost never<br>(n=21,276) |  |
|  | n | % | n | % | n | % | n | % |
| Age, mean (standard deviation) | 49.6 | (10.0) | 48.7 | (10.3) | 47.6 | (10.4) | 46.5 | (10.6) |
| Sex, male | 1,576 | 56.5 | 860 | 58.2 | 925 | 62.0 | 10,453 | 49.1 |
| Marital status |  |  |  |  |  |  |  |  |
| Married | 1,406 | 50.4 | 915 | 61.9 | 959 | 64.2 | 11,749 | 55.2 |
| Divorced/Widowed | 284 | 10.2 | 114 | 7.7 | 106 | 7.1 | 2,339 | 11.0 |
| Unmarried | 1,100 | 39.4 | 448 | 30.3 | 428 | 28.7 | 7,188 | 33.8 |
| Job type |  |  |  |  |  |  |  |  |
| Mainly desk work | 2,042 | 73.2 | 1,058 | 71.6 | 952 | 63.8 | 9,416 | 44.3 |
| Mainly involving interpersonal communication | 495 | 17.7 | 333 | 22.5 | 391 | 26.2 | 5,708 | 26.8 |
| Mainly physical work | 253 | 9.1 | 86 | 5.8 | 150 | 10.0 | 6,152 | 28.9 |
| Annual equivalent household income (JPY) |  |  |  |  |  |  |  |  |
| < 2,500,000 | 751 | 26.9 | 207 | 14.0 | 189 | 12.7 | 4,563 | 21.4 |
| 2,500,000–3,750,000 | 643 | 23.0 | 297 | 20.1 | 305 | 20.4 | 6,305 | 29.6 |
| 3,750,000–5,000,000 | 576 | 20.6 | 362 | 24.5 | 371 | 24.8 | 5,316 | 25.0 |
| > 5,000,000 | 820 | 29.4 | 611 | 41.4 | 628 | 42.1 | 5,092 | 23.9 |
| Education |  |  |  |  |  |  |  |  |
| Junior high school | 31 | 1.1 | 13 | 0.9 | 6 | 0.4 | 318 | 1.5 |
| High school | 507 | 18.2 | 198 | 13.4 | 238 | 15.9 | 6,010 | 28.2 |
| Vocational school, junior college, university, or graduate school | 2,252 | 80.7 | 1,266 | 85.7 | 1,249 | 83.7 | 14,948 | 70.3 |
| Current smoker | 693 | 24.8 | 406 | 27.5 | 441 | 29.5 | 5,464 | 25.7 |
| Alcohol consumption |  |  |  |  |  |  |  |  |
| 6 or more days a week | 658 | 23.6 | 360 | 24.4 | 341 | 22.8 | 4,315 | 20.3 |
| 4 or 5 days a week | 234 | 8.4 | 165 | 11.2 | 163 | 10.9 | 1,515 | 7.1 |
| 2 or 3 days a week | 316 | 11.3 | 195 | 13.2 | 241 | 16.1 | 2,514 | 11.8 |
| Within 1 day a week | 445 | 15.9 | 262 | 17.7 | 281 | 18.8 | 3,559 | 16.7 |
| Almost never | 1,137 | 40.8 | 495 | 33.5 | 467 | 31.3 | 9,373 | 44.1 |
| Number of employees |  |  |  |  |  |  |  |  |
| 1–9 | 1,670 | 59.9 | 343 | 23.2 | 275 | 18.4 | 3,877 | 18.2 |
| 10–99 | 228 | 8.2 | 230 | 15.6 | 245 | 16.4 | 6,237 | 29.3 |
| 100–999 | 321 | 11.5 | 335 | 22.7 | 375 | 25.1 | 6,122 | 28.8 |
| >1000 | 571 | 20.5 | 569 | 38.5 | 598 | 40.1 | 5,040 | 23.7 |
| Work functioning impairment scale (Wfun) score |  |  |  |  |  |  |  |  |
| 7–20 | 2,163 | 77.5 | 1,134 | 76.8 | 1,154 | 77.3 | 16,794 | 78.9 |
| 21–35 | 627 | 22.5 | 343 | 23.2 | 339 | 22.7 | 4,482 | 21.1 |
| Teleworking preference |  |  |  |  |  |  |  |  |
| I would like to telework as much as possible | 1,789 | 64.1 | 463 | 31.3 | 259 | 17.3 | 2,196 | 10.3 |
| I prefer teleworking | 409 | 14.7 | 440 | 29.8 | 376 | 25.2 | 2,666 | 12.5 |
| I don't mind either | 436 | 15.6 | 376 | 25.5 | 483 | 32.4 | 6,637 | 31.2 |
| I prefer to work in the workplace | 98 | 3.5 | 154 | 10.4 | 272 | 18.2 | 2,845 | 13.4 |
| I prefer to work in the workplace as much as possible | 58 | 2.1 | 44 | 3.0 | 103 | 6.9 | 6,932 | 32.6 |

Table 2 shows the association of work functioning impairment with teleworking preference and frequency. In the age- and sex-adjusted model, strong preference for teleworking was associated with work functioning impairment (odds ratio [OR]: 2.08, 95% confidence interval [CI]: 1.90–2.28). The same trend was observed for Model 1, which was adjusted for socioeconomic and lifestyle factors (OR: 2.20, 95% CI: 2.01–2.42) and Model 2, which was adjusted for socioeconomic and lifestyle factors and teleworking frequency (OR: 2.27, 95% CI: 2.05–2.51), with participants who prefer teleworking as much as possible associated with work functioning impairment. In the age- and sex-adjusted model, the OR of work functioning impairment was significantly higher among participants who teleworked 4 or more days a week than workers who almost never teleworked (OR: 1.18, 95% CI: 1.07–1.3). In Model 1, adjusted for socioeconomic and lifestyle factors, the OR of work functioning impairment was significantly higher among employees who teleworked 4 or more days a week (OR: 1.32, 95% CI: 1.18–1.46) and those who teleworked 2 or 3 days a week (OR: 1.28, 95% CI: 1.12–1.46) compared with those who almost never teleworked. In Model 2, adjusted for teleworking preference in addition to socioeconomic and lifestyle factors, the ORs of work functioning impairment were no longer significantly different among workers who teleworked 4 or more days a week (OR: 0.90, 95% CI: 0.80–1.01) and those who teleworked 2 or 3 days a week (OR: 1.01, 95% CI: 0.88–1.16).

**Table 2.**
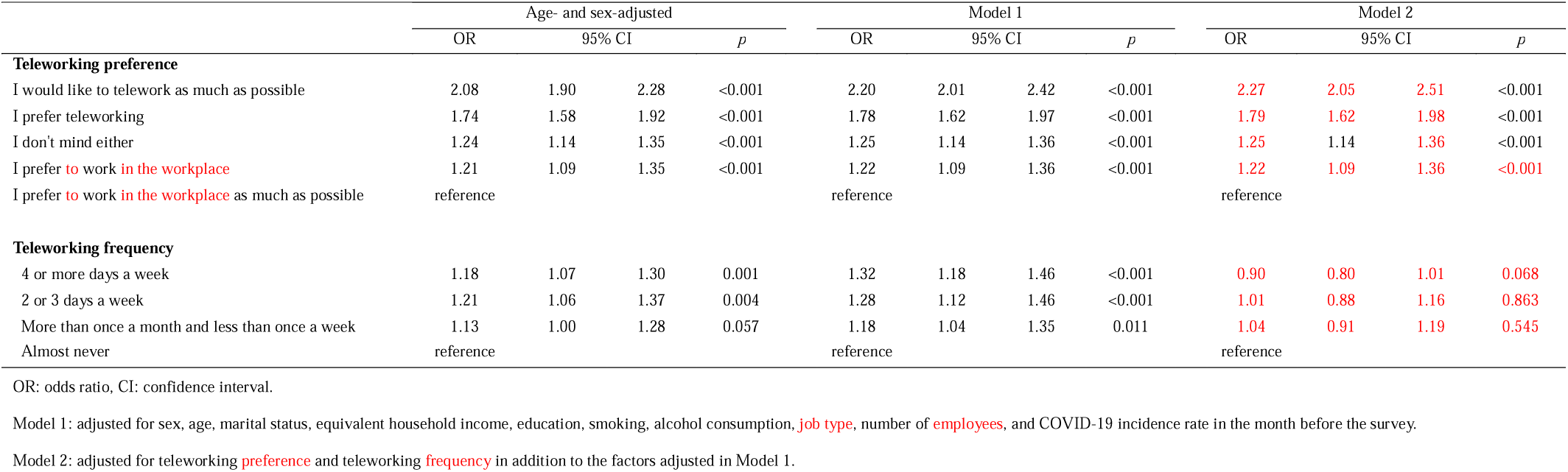
Odds ratio of work functioning impairment associated with teleworking preference and frequency

|  | Age- and sex-adjusted |  |  |  | Model 1 |  |  |  | Model 2 |  |  |  |
| --- | --- | --- | --- | --- | --- | --- | --- | --- | --- | --- | --- | --- |
|  | OR | 95% CI |  | *p* | OR | 95% CI |  | *p* | OR | 95% CI |  | *p* |
| **Teleworking preference** |  |  |  |  |  |  |  |  |  |  |  |  |
| I would like to telework as much as possible | 2.08 | 1.90 | 2.28 | <0.001 | 2.20 | 2.01 | 2.42 | <0.001 | 2.27 | 2.05 | 2.51 | <0.001 |
| I prefer teleworking | 1.74 | 1.58 | 1.92 | <0.001 | 1.78 | 1.62 | 1.97 | <0.001 | 1.79 | 1.62 | 1.98 | <0.001 |
| I don't mind either | 1.24 | 1.14 | 1.35 | <0.001 | 1.25 | 1.14 | 1.36 | <0.001 | 1.25 | 1.14 | 1.36 | <0.001 |
| I prefer to work in the workplace | 1.21 | 1.09 | 1.35 | <0.001 | 1.22 | 1.09 | 1.36 | <0.001 | 1.22 | 1.09 | 1.36 | <0.001 |
| I prefer to work in the workplace as much as possible | reference |  |  |  | reference |  |  |  | reference |  |  |  |
| **Teleworking frequency** |  |  |  |  |  |  |  |  |  |  |  |  |
| 4 or more days a week | 1.18 | 1.07 | 1.30 | 0.001 | 1.32 | 1.18 | 1.46 | <0.001 | 0.90 | 0.80 | 1.01 | 0.068 |
| 2 or 3 days a week | 1.21 | 1.06 | 1.37 | 0.004 | 1.28 | 1.12 | 1.46 | <0.001 | 1.01 | 0.88 | 1.16 | 0.863 |
| More than once a month and less than once a week | 1.13 | 1.00 | 1.28 | 0.057 | 1.18 | 1.04 | 1.35 | 0.011 | 1.04 | 0.91 | 1.19 | 0.545 |
| Almost never | reference |  |  |  | reference |  |  |  | reference |  |  |  |

OR: odds ratio, CI: confidence interval.
Model 1: adjusted for sex, age, marital status, equivalent household income, education, smoking, alcohol consumption, **job type**, number of **employees**, and COVID-19 incidence rate in the month before the survey.
Model 2: adjusted for teleworking **preference** and teleworking **frequency** in addition to the factors adjusted in Model 1.

## Discussion

In this study, we found an association between teleworking frequency and increased work functioning impairment. Additionally, workers who preferred teleworking experienced increased work functioning impairment. When teleworking preference and frequency were adjusted for, only teleworking preference was associated with work functioning impairment.

A remarkable finding of our study was that workers who preferred to telework had greater work functioning impairment. The preference for teleworking could be based on difficulties with working in the workplace owing to ill health or work–family conflicts. Previous studies have reported that workers who prefer telework choose teleworking when feeling unwell because this allows them to work at their own pace and freely adjust their break time.^26,27^ Other previous studies state that female employees with children choose to telework but are not highly satisfied with telework.^28,29^ Attention to the background related to teleworking preferences is important to support teleworkers with severe work functioning impairment.

The current study showed that frequent teleworking was associated with work functioning impairment. One important mechanism for this association could be that individuals with health problems are more likely to choose to telework. ^27,30^ In this study, when teleworking preference and teleworking frequency were adjusted, a teleworking preference was associated with work functioning impairment whereas the teleworking frequency was no longer associated with work functioning impairment. Some studies have suggested that teleworking is associated with poor health conditions, such as depression and musculoskeletal pain.^31,32^ Key to detecting signs of work functioning impairment would be to follow up on the health conditions of employees who frequently telework.

This study has several public health implications. First, governments should create a system that enables teleworkers to continue working with reduced work function. Early detection of work functioning impairment is more difficult in telework than in the workplace where there is face-to-face interaction. Strategies to check work function are necessary, even for teleworkers. For example, a checklist to verify the work function of teleworkers should be developed and promoted for regular use. Second, companies should pay attention to background factors that reduce the work function of teleworkers. Identifying needed support for teleworkers will prevent impaired work function. Third, companies should assess the risk of absence from work for individual workers and provide appropriate work accommodations in cooperation with occupational health staff. Occupational health staff should have online meetings with teleworkers to check for any health problems and provide information on health management when teleworking.

Our study has several limitations: First, the self-reported data used in our study may be inaccurate. Self-reporting may lead to systematic errors, which may reduce or increase the significance of the differences. Second, this was a cross-sectional study, and it is not possible to determine causality. For example, we cannot determine whether there is a causal relationship between teleworking frequency and teleworking preferences.

In conclusion, the current study findings suggest that a teleworking preference is associated with work functioning impairment and one factor that increases the teleworking frequency. This result may be explained by workers who have difficulties with working in the workplace owing to ill health or work–family conflicts prefer to telework. Employees who telework frequently should be carefully monitored to determine whether the ability to perform their job is impaired. Investigating the background regarding the choice to telework could provide clues to understanding the support needed by teleworkers, which will help to prevent health problems and lead to increased productivity among these workers.

## Data Availability

Data not available due to ethical restrictions.

## Notes

### Competing Interest Statement

Dr. Fujino reports personal fees from Sompo Health Support Inc. for the copyright of WFun. The other authors have no conflicts of interest to declare regarding this study.

### Funding Statement

a research grant from the University of Occupational and Environmental Health, Japan (no grant number); Japanese Ministry of Health, Labour and Welfare (grants H30-josei-ippan-002, H30-roudou-ippan-007, 19JA1004, 20JA1006, 210301-1, and 20HB1004); Anshin Zaidan (no grant number), the Collabo-Health Study Group (no grant number), and Hitachi Systems, Ltd. (no grant number) and scholarship donations from Chugai Pharmaceutical Co., Ltd. (no grant number).

### Author Declarations

This study was conducted with approval from the Ethics Committee of University of Occupational and Environmental Health, Japan (Approval No. R2-079).

### Summary of Updates

We only changed the "competing interests" part.

